# “Circumstantial Determinants”: An Efficient Approach to Reaching People in Need of HIV Prevention?

**DOI:** 10.64898/2026.06.18.26355534

**Authors:** S Bagnay, S Gregson, M Skovdal, R Maswera, L Moorhouse, G Ncube, B Tsenesa, P Mandizvidza, M Pickles, GP Garnett, O Mugurungi, C Nyamukapa

**Affiliations:** Imperial College London, Department of Infectious Disease Epidemiology, London, England; Biomedical Research and Training Institute, Harare, Zimbabwe; University of Copenhagen, Department of Public Health, Copenhagen, Denmark; Zimbabwe Ministry of Health and Child Care, Harare, Zimbabwe; London School of Hygiene and Tropical Medicine, Faculty of Public Health and Policy, London, England

**Keywords:** HIV prevention, HIV testing, targeted prevention, Circumstantial Determinants, HIV prevention cascade

## Abstract

HIV prevention and testing programmes primarily reach people who self-refer or attend routine health services. Higher-risk individuals are missed if they are healthy, underestimate their risk of infection or under-report sexual risk-behaviours. We assess a new approach to address limitations in existing programmes by targeting HIV services on “Circumstantial Determinants” (CDs) of HIV risk – the social circumstances, settings, and norms associated with behaviours that increase risk of HIV acquisition.

Data on potential CDs and sexual behaviour were collected in a population survey in Zimbabwe in 2018/19 (N=9141). HIV-negative individuals reporting ≥1 sexual risk-behaviours were defined as the’priority population’ for HIV prevention. For each sex, six circumstantial determinants were associated with being in the priority population (aOR≥1.30; p≤0.01). Reach and efficiency of CDs (and combinations) were calculated; ROC curve algorithms evaluated their ability to identify priority population membership; and HIV prevention condom cascades were compared between CD-defined priority population subgroups.

Example findings include that targeting men at bars and beerhalls could reach 48.5% of the priority population and 25.1% of lower-risk men. These percentages increase to 77.1% and 53.7% if men with poor mental health, no religious affiliation, negative social capital, or living on agricultural estates are also targeted. Targeting women with poor mental health could reach 32.0% of the priority population and 21.3% of lower-risk women. Targeting additional circumstantial determinants increases these percentages to 54.1% and 37.5%, respectively. Cascade barriers to condom use differed between CD-defined subgroups. The Circumstantial Determinants approach demonstrates proof-of-concept potential to strengthen HIV prevention services.

## Introduction

Despite good progress made in scaling up antiretroviral treatment (ART) and prevention services, 1.3 million new people are estimated to have become infected in 2024 (UNAIDS, 2025). Achieving further reductions in HIV incidence through primary prevention - and associated HIV testing for methods that require this - could be made more efficient and effective if we include more at-risk individuals in programmes while minimising the total number of people included.

For HIV prevention, given limited resources, HIV testing must target those at higher risk of acquiring HIV infection. Many studies have looked at risk factors for acquiring HIV. After controlling for confounding, relatively few causal risk factors have been identified (Chen et al., 2007). However, in designing programmes and targeting service delivery to maximise impact and efficiency, we are interested in variables along the causal pathway and correlated with HIV as well as those that directly lead to acquisition. Risk scores have been developed and studied to screen people for HIV risk and prioritise service delivery. These scores have focused on an individual’s biological and behavioural variables, but their performance has generally been poor (Jia et al., 2022). This limitation reflects a broader problem: biological and behavioural risks do not arise in isolation, but they are embedded in and shaped by the socio-cultural and economic environments that create the context and circumstances where HIV transmission can occur. By understanding these circumstances, we may be better able to focus HIV services.

A number of approaches currently used in national HIV control programmes focus on individuals’ biological and behavioural characteristics. Principal among these is routine screening of individuals attending healthcare services, followed by testing of those who report current or recent HIV risk behaviours or sexually transmitted infections, or who are pregnant. Strategies such as HIV testing at standalone centres, index case testing (Katbi et al., 2018) and HIV self-testing (Johnson et al., 2014) are often used to reach those at risk who are not seen regularly in healthcare services. However, from an HIV prevention perspective, there are important limitations in these approaches. These include that the HIV risk screening procedures used in routine testing services rely on self-reports that often understate socially-proscribed high risk sexual behaviours; that use of standalone testing centres and HIV self-test kits can be limited due to low HIV risk perception (Schaefer et al., 2020); and that reports of past and present behaviours may not be predictive of future risk behaviours (Moorhouse et al., 2024).

Some targeting approaches already exist that avoid these limitations. These include the PLACE method (Weir et al., 2003), which pinpoints specific sites or’hot spots’ with high rates of new sexual partnership formation (Chang et al., 2016; Popoola et al., 2024) and targets these with HIV services, and, relatedly, interventions in bars and beerhalls and with identifiable population sub-groups with high levels of sexual partnership formation (e.g. female sex workers and male factory workers) (Wilson et al., 1990). A key aspect of these approaches is that, rather than selecting or screening for individual-level characteristics previously found to be associated with HIV risk, they focus primarily on the social, cultural, and economic circumstances within which the chances that individuals will engage in HIV risk behaviours typically increase. Furthermore, these circumstances include places and venues that can be visited, where, for example, HIV testing services can be offered or at least actively promoted to people who attend, without the need to conduct screening interviews. However, not all new infections occur in these locations and groups, and other social circumstances and experiences have also been found to be associated with higher levels of HIV infection in some populations – for example, women experiencing intimate partner violence (Wagman et al., 2015) and people experiencing mental health problems (Kelebie et al., 2025). Therefore, it may be useful to consider whether broadening this approach to include other social circumstances and experiences might be an effective and practical way to increase use of HIV prevention methods among those who need them most.

In this paper, we introduce and explore the utility of a novel targeting approach for HIV testing and prevention based on a wider range of social circumstances and a potentially unifying concept of “Circumstantial Determinants”, which we define as:

> *“Circumstantial determinants of HIV infection refer to the spatial settings, social interactions, life situations or events, socio-cultural norms, and individual predispositions that either on their own or in combination, give rise to specific circumstances that increase exposure to the risks of acquiring an illness or injury.”*

Thus, the Circumstantial Determinants (CD) approach focuses on social contexts – that comprise a mix of social circumstances, settings and norms, and alternative combinations of these contexts – and examines if it can be used to identify individuals currently overlooked in existing mainstream approaches. Assessment of this approach to identifying HIV risk requires two main steps. First, variables well known to be directly associated with HIV acquisition (such as multiple sex partners) are used to identify a priority population. Second, social, spatial, attitudinal and less proximal behavioural variables are identified that predict whether individuals fall within this priority population. The likelihood that someone in a priority population will acquire HIV – which will determine the cost-effectiveness of HIV interventions – is a function of HIV prevalence in the partner pool and coverage of HIV interventions. Any targeted approach will be more efficient than universal delivery, provided that the cost of identifying and reaching the priority population does not exceed the savings achieved by targeting a smaller population (Garnett et al., 2017).

As a proof-of-concept, this study tests the following two hypotheses: (1) that incorporating circumstantial determinants within the design of HIV prevention programmes can better target those at risk of HIV acquisition, and (2) that outcomes across the HIV prevention cascade depend on the circumstances that create this risk.

## Methods

### Analytic approach

This study used four sequential steps to test the CD approach for HIV prevention targeting.

*Hypothesis 1*

1) Identification of the circumstantial determinants (CDs) and combinations of CDs that are associated with engaging in sexual behaviours that increase the risk of acquiring HIV infection, and measure and compare the strengths of these associations (Note: henceforth in this paper, HIV-negative individuals who engage in these sexual behaviours are referred to as the’priority population’ for use of HIV prevention methods);
2) Assessment of the coverage and relative efficiency of targeting different CDs, individually and in combination, through estimation and comparison of the proportions of the priority population reached and the general population targeted for each possible combination of CDs; and
3) Development of a ROC curve algorithm for the HIV-negative population, based on the selected CDs, to predict high-risk sexual behaviour, and evaluation of the algorithm’s ability to identify individuals in the priority population for HIV prevention; and

*Hypothesis 2*

4) Measurement of the HIV prevention condom cascades for the sub-populations within the priority population that could potentially be reached by targeting each CD, and identification of differences and similarities between these cascades.

### Data

Data were taken from the seventh round of the Manicaland General Population Open-Cohort survey (Manicaland survey), conducted between July 2018 and December 2019 (www.manicalandhivproject.org). Eight sites were included in the survey, representing five of Manicaland’s main socioeconomic strata: urban areas, rural areas, agricultural estates, rural business centres, and major transport routes. As in previous survey rounds (Gregson et al., 2017), individuals aged 15 years and above resident in a two-thirds random sample of households enumerated in an updated census were eligible for interview in the 2018/19 round. To provide additional data for trials of HIV prevention interventions in young people, females aged 15-24 and males aged 15-29 resident in the remaining one-third of households were also treated as eligible for the individual survey. On this basis, 10,691 household members were eligible to participate of which 9,803 (91.7%) provided informed consent, and 9,141 (5,321 females and 3,820 males) fell within the eligible age-range for this study (15-65 years). Data collected included information on sociodemographic characteristics, sexual risk behaviours (SRBs) for HIV acquisition, mental health, social norms, and use of HIV services.

The data were collected in face-to-face interviews lasting approximately 1 to 1.5 hours. Interviews were conducted in either Shona or English, depending on the participant’s preference. Informal confidential voting interview (ICVI) methods were used to reduce social desirability bias in data on sexual behaviour (Gregson et al., 2002). Dried blood spot samples (DBS) were collected for HIV testing.

### Study measures

The *general population* includes all males and females aged 15-65, regardless of their HIV status. The *priority population* is limited to HIV-negative sexually-active individuals aged 15-54 at higher risk of acquiring HIV, who could, therefore, benefit from HIV prevention methods. The 15-54 range is used in comparable studies and reflects the decline in risk-behaviours after age 54 (Negin et al., 2016; Bagnay et al., 2025). In the study, HIV-negative participants reporting one or more of the following behaviours – previously found to be associated with HIV (Lopman et al., 2008) – were considered to be at higher risk of acquiring HIV and therefore part of the priority population for HIV prevention methods: (1) multiple sexual partners in the past 12 months; (2) concurrent sexual partners (currently); (3) transactional sex with any of the last three sexual partners in the past month; (4) one or more non-regular sexual partner(s) in the past 12 months; and, for women, (5) age-disparate relationships (partner 10+ years older) with any of the last three sexual partners (see Bagnay et al., 2026 (Section 1) for more details).

A preliminary review of literature was carried out to identify circumstantial factors previously found to be associated with increased sexual risk behaviour that potentially could be used to identify individuals not reached using exclusively individual-based targeting strategies (Bagnay et al., 2026, Section 2). More details on the rationale behind the selected CDs and the categorisation can be found in Bagnay et al. (2026), Section 2. Circumstantial determinants investigated in this study are:

(1) Residence in or close to a large-scale agricultural estate compound, rural business centre, or major transport route (Weir et al., 2003);
(2) Religious beliefs (Manzou et al., 2014; Shaw & El-Bassel, 2014);
(3) Negative social capital (Gregson et al., 2004);
(4) Regular visits to bars or beerhalls (Fritz et al., 2011; Lewis et al., 2005);
(5) Women living in poverty whose partners spend extended periods away from home (Lurie et al., 2003; Nicholas et al., 2016);
(6) Young people of school age who are out of school (Gregson et al., 2001; Nutakor et al., 2023);
(7) Exposure to intimate partner violence (IPV) (Cordeiro et al., 2024; Townsend et al., 2011; Wagman et al., 2015);
(8) Experiencing poor mental health (Kelebie et al., 2025; Tlhajoane et al., 2018);
(9) Holding masculine social norms associated with sexual risk behaviour (Bagnay et al., 2025; Sileo et al., 2019), and
(10) Having access to a mode of transport (Weir et al., 2003; Lurie et al., 2003).

This generated ten CDs for females and eight for males that have previously been found to increase the likelihood that individuals engage in sexual risk behaviours for HIV acquisition.

## Statistical analysis

Descriptive statistics were used to describe the sociodemographic characteristics of the study population and the frequencies of these ten CDs. Weighted percentages were used to account for the oversampling of young people.

### Selecting circumstantial determinants for inclusion in models to identify priority populations

Age-adjusted odds ratios (AORs), 95% confidence intervals (95% CIs), and *p*-values were calculated for each category using multiple logistic regression to estimate the likelihood of exposure within the priority population compared to the general population. An arbitrary threshold was set at an OR≥1.30 and *p*≤0.01. Categories reaching this threshold were selected for further analysis, as exposure was considered to be both materially and significantly higher in the priority population. Weighted risk ratios (RRs), to account for the over-sampling of young people, were calculated for these categories. The CD’adolescents that dropped out of school’ was excluded from further analysis due to the small number of participants. These selected circumstantial determinants were included in further analyses.

*Combinations of selected circumstantial determinants* were compared. For each combination, the percentage of the general population that would be targeted (with HIV prevention programmes) and the percentage of the priority population that could be reached when targeting were calculated, taking overlaps between CDs into account (the’AND/OR’, and’AND’ approaches). Venn diagrams were created to visualise overlaps. AORs and *p*-values were calculated for overlaps between combinations of up to three CDs using multiple logistic regression analysis, measuring the occurrence of those combinations within the priority population compared to within the general population. RRs were calculated to compare the percentage of the general population that would be targeted with the percentage of the priority population that could be reached. Heatmaps were created to visualise RRs for the ‘and/or’ approach. For several combinations with the highest RRs, practicality ratings were assigned after deliberation within the research team. These practicality ratings provide insight into how we perceive the difficulty of targeting these combinations of CDs.

*Receiver Operating Characteristic (ROC) curve algorithms* were developed and tested for the HIV-negative population to evaluate the CD approach for prevention programme development and its ability to accurately classify individuals with SRBs based on exposure to the CDs. The first algorithm employed a stepwise approach, using the Area Under the Curve (AUC) as a measure of overall performance to balance sensitivity and specificity. In this approach, the test algorithm followed the order of CDs as established in the training algorithm. The second algorithm also optimised AUC but differed in that it allowed flexibility in the sequence of introducing CDs. At each step, the CD yielding the greatest incremental increase in AUC was chosen, regardless of whether this order matched the training dataset. The third algorithm employed a similar approach to the first algorithm but focused on sensitivity to maximise the identification of individuals with SRBs instead of AUC. Like the first algorithm, the test algorithm followed the order established in the training algorithm. The fourth algorithm was similar to the second, but it focused on maximising sensitivity rather than AUC. By examining these four different algorithms, we aimed to include different perspectives. HIV-positive individuals and those with unknown HIV status were excluded from these algorithms. The outcome variable was defined as being in the priority population in need of HIV prevention. Two datasets were created using all HIV-negative males and females aged 15-65. Training datasets were used to develop the algorithm, while the test datasets evaluated the model established in the training dataset. 75.0% of the HIV-negative population was randomly assigned to the training dataset (males: N=2548, females: N=3381) and 25.0% to the test dataset (males: N=796, females: N=1099). Each CD was assessed for the strength of its ability to correctly classify the outcome variable; sensitivity (true positives (TP)/TP + false negatives (FN)), specificity (true negatives (TN)/TN+ false positives (FP), OR, likelihood ratio (LR+), and AUC were calculated. A stepwise iterative process was used to add CDs, starting with the CD with the highest AUC or sensitivity for classifying individuals as being in the priority population. For each step of the algorithm, the positive predictive value (PPV) (TP/TP+FP), the negative predictive value (TN/TN+FN), and the accuracy (TP+TN/total) were calculated. ROC curves for the first and second algorithms were used to visualise the trade-offs between decreasing specificity and increasing sensitivity for the first algorithm.

*HIV prevention male condom cascades (HPCs)* (Schaefer et al., 2019) were populated for males and females within the priority population across selected CDs to assess whether and how gaps in recent condom use with non-regular sexual partners differ between individuals exposed through different CDs. The HPC is a’conditional cascade’, meaning that each additional step is conditional on the preceding step. Male condoms were chosen as a primary prevention method due to their high accessibility and broad reach in Zimbabwe (Zimbabwe National Statistics Agency and ICF, 2025). Following the previous steps, two CDs per sex were selected based on sample size, practicality ratings, visualisation gaps in the test HPCs, and the ROC curve algorithm. Those who reported not having a non-regular partner were excluded from the HPC analysis. Differences between cascades were identified through visualisation and logistic regression analysis. When comparing CDs statistically, the overlapping population was excluded from the analysis. However, these individuals were included in the HPCs to avoid losing valuable data.

All analyses were disaggregated by sex (male/female) and conducted using STATA version 17.0. ROC curves were generated in Excel. HPCs were created using Tableau version 2024.2.1, and Heatmaps were created in RStudio version 2025.05.1.

## Results

### Circumstances associated with being in the priority population for HIV prevention in Manicaland, east Zimbabwe

#### Identifying circumstantial determinants in Manicaland

Table 1 presents the proportions of the priority and general populations within each CD category. Categories identified for further analysis, based on the odds ratio and probability differences, are shown in italics. For males, 15.5% of the general population were part of the priority population. For females, this percentage was 18.8%.

**Table 1:**
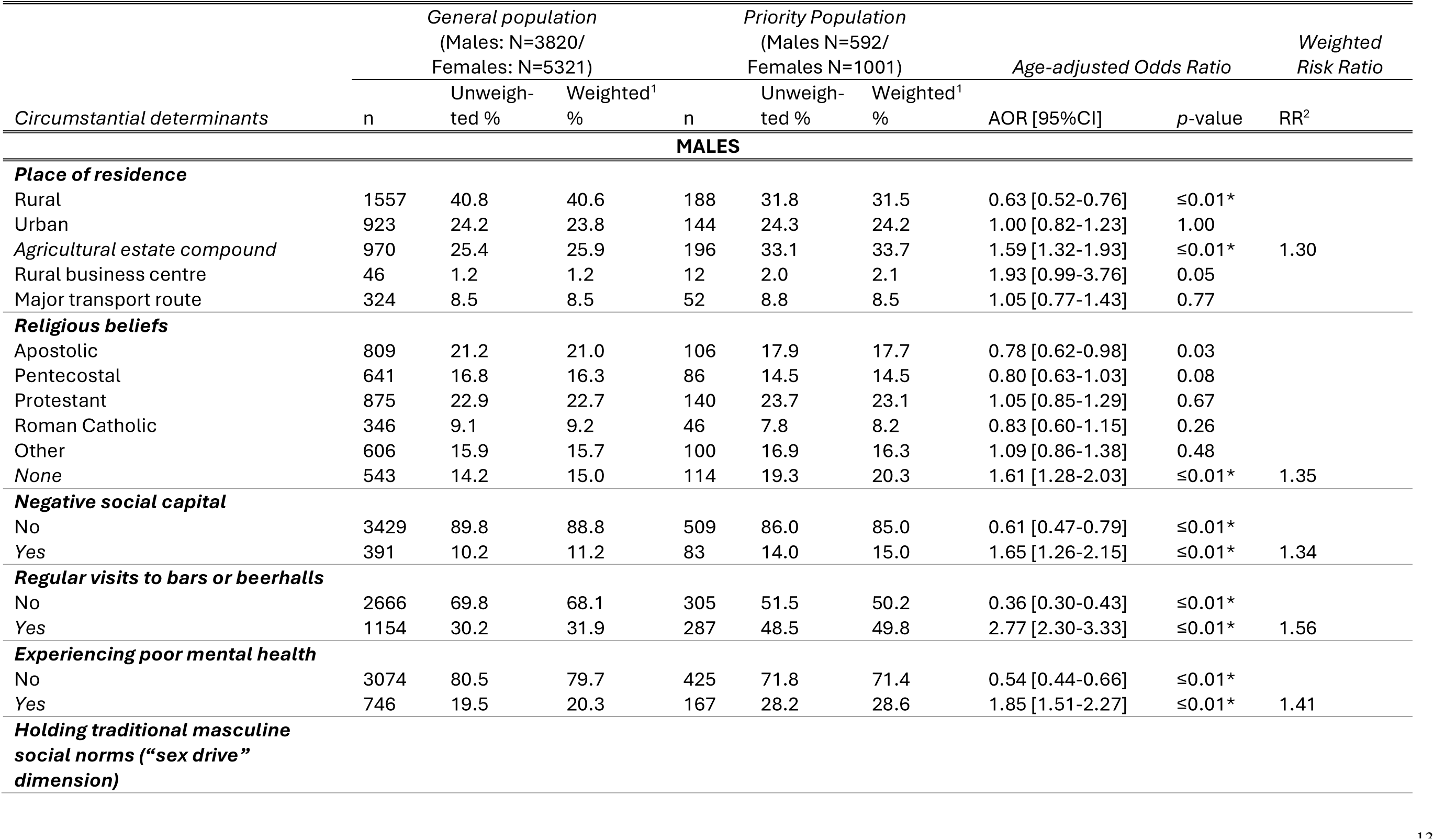
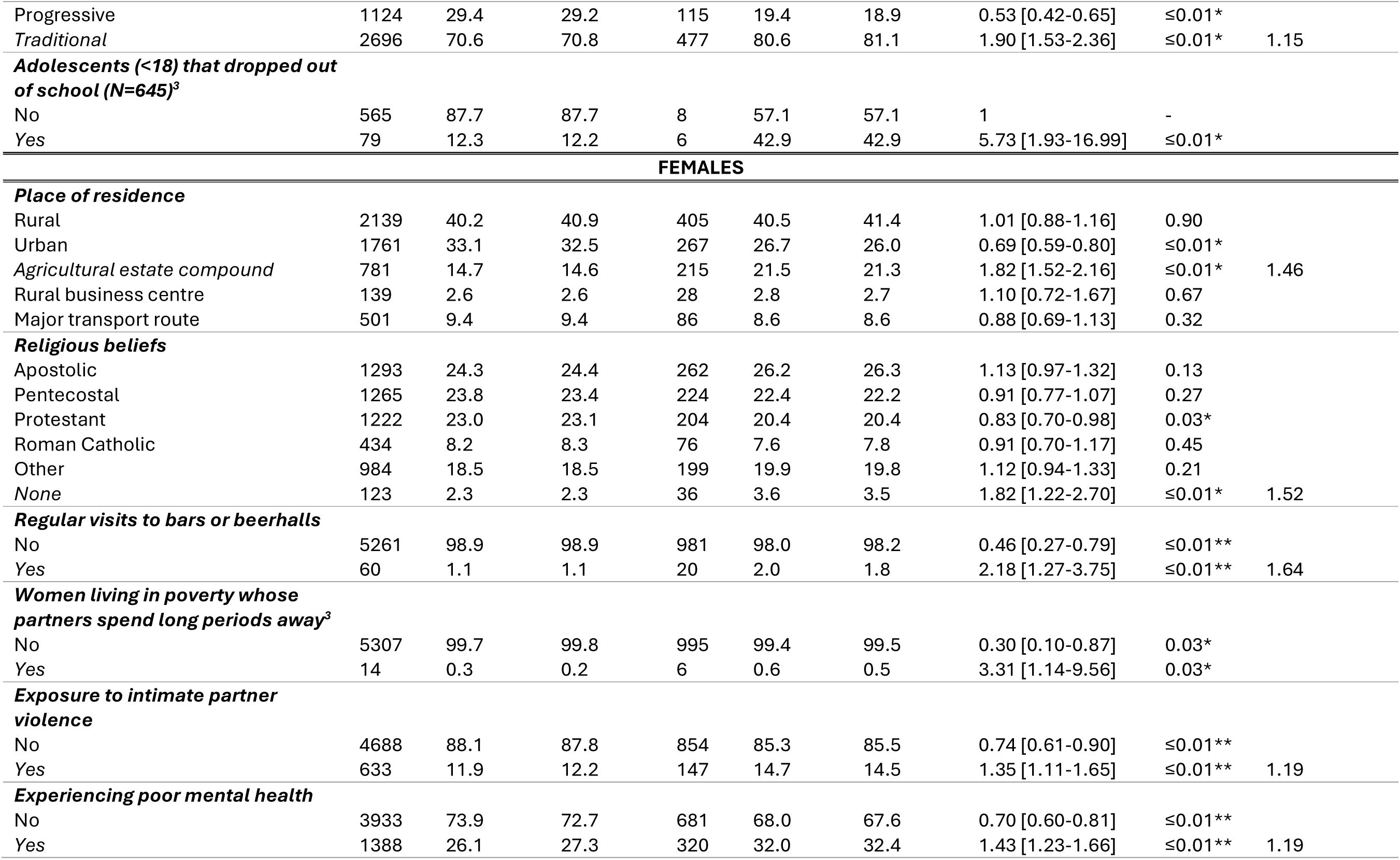
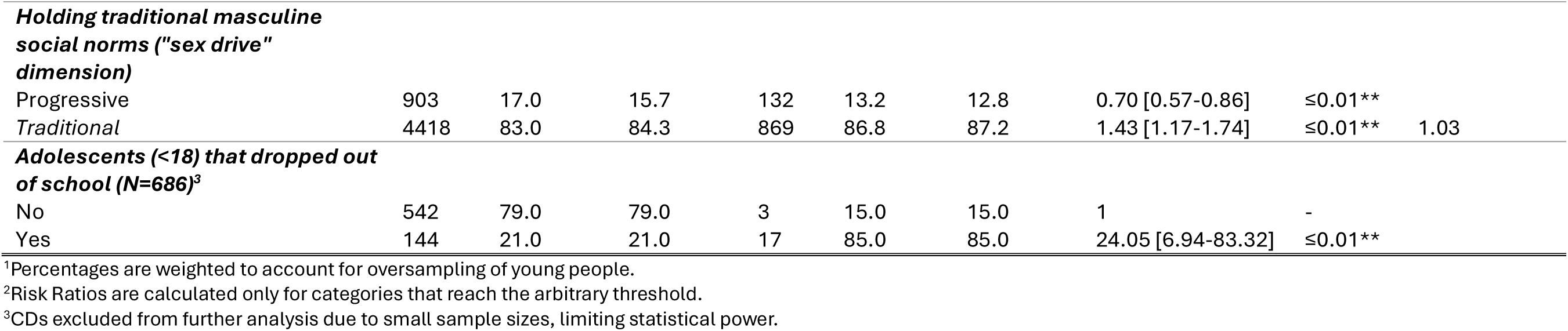
Selected circumstantial determinants (CDs) reaching an arbitrary threshold of OR≥1.30 and *p*≤0.01 for the general population aged 15-65, and the priority population consisting of HIV-negative individuals aged 15-54 with one or more sexual risk behaviours in the past 12 months in Manicaland, Zimbabwe. The age-adjusted odds ratios and weighted risk ratios indicate the difference in risk between the priority and general populations. Asterisks (*) highlight statistically significant differences (*p*≤0.05).

Among males, the qualifying CDs were: (1) residence in an agricultural estate compound; (2) no religious affiliation; (3) negative social capital; (4) regular visits to bars or beer halls; (5) poor mental health, and (6) conservative masculine social norms (“sex drive” dimension). For example, as shown in Table 1, men in the priority population had 2.77 times higher odds of regularly visiting bars or beerhalls, and their associated risk of regular visits was found to be 56.0% higher compared to men in the general population. For females, the qualifying CDs were: (1) residence on agricultural estate compounds; (2) no religious affiliation; (3) regular visits to bars and beerhalls; (4) experience of intimate partner violence; (5) poor mental health; and (6) conservative masculine norms (“sex drive” dimension) (Table 1). Most of the selected CDs showed lower coverage in females than in males.

*Combinations of circumstantial determinants* reaching the qualifying threshold were compared and analysed (Bagnay et al., 2026, Section 6). A selection of these combinations of determinants with the highest RRs is shown in Table 2 (illustrative examples). The results show a trend of diminishing RRs for combinations that include increasing numbers of CDs.

**Table 2:**
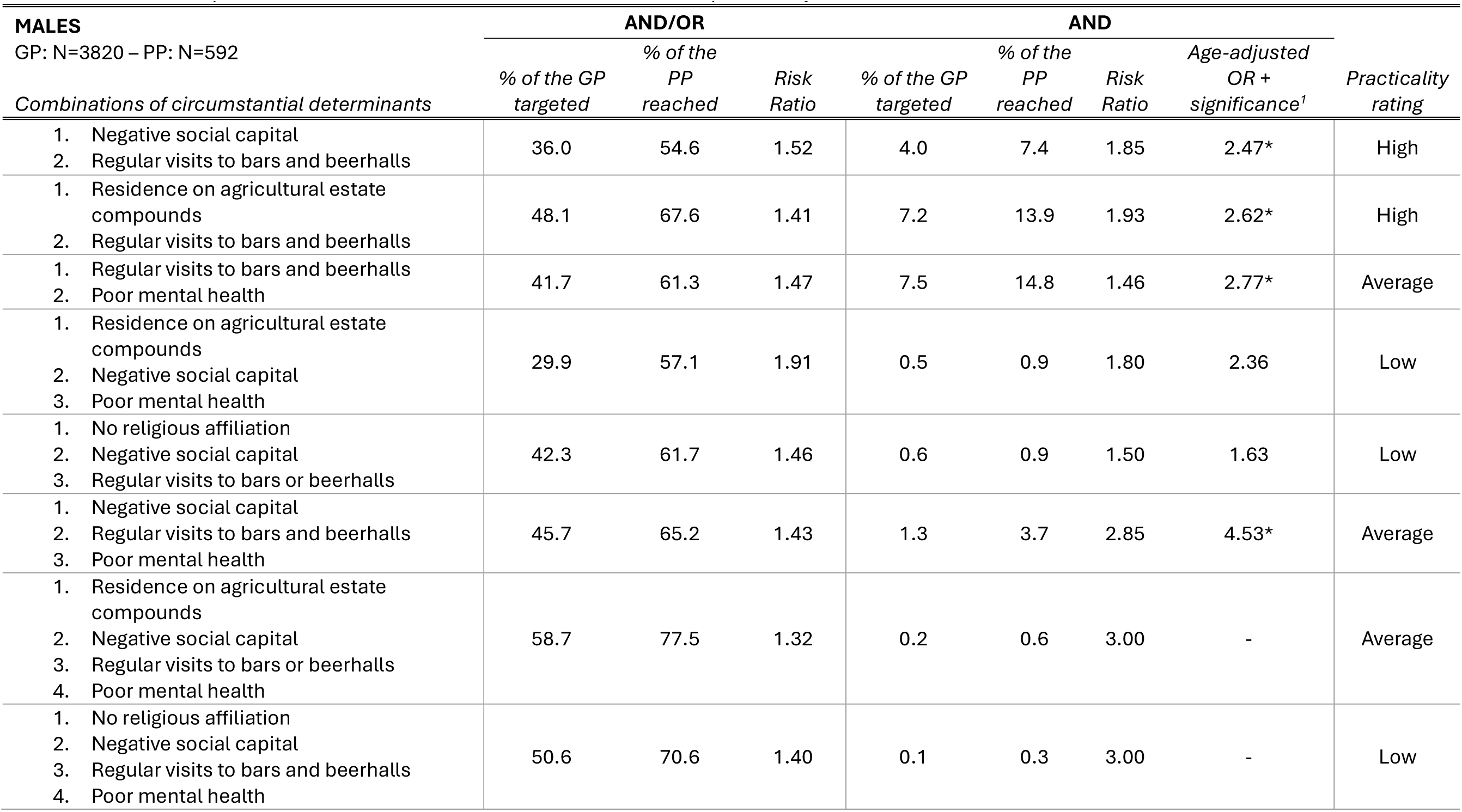
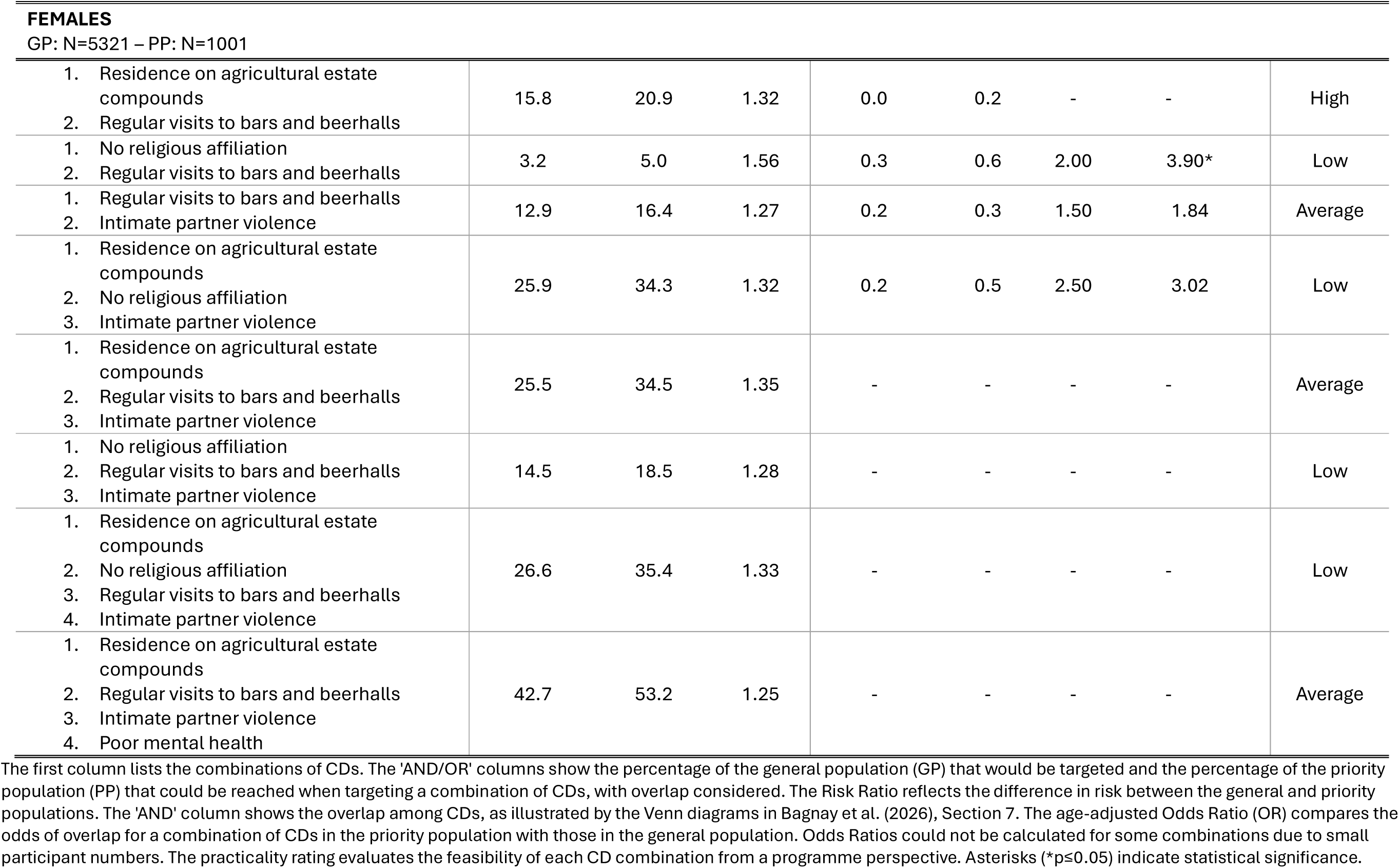
A selection of combinations of circumstantial determinants reaching an arbitrary threshold of OR≥1.30 and *p*≤0.01 with high-risk ratios (RR) for the’AND/OR’ and’AND’ (only where CDs are overlapping) combinations with practicality ratings, using data from the Manicaland general-population survey 2018-2019. Due to limited space, not all high-RR combinations are shown here (see Bagnay et al., 2026, Section 6, for full tables). These combinations were selected by our team to provide diverse, illustrative examples and to demonstrate internal assessments of their practicality.

When targeting males with negative social capital and those who regularly visit bars and beerhalls (using the AND/OR approach), 36.0% of the general population must be targeted to reach 54.6% of the priority population (Table 2). The associated RR is 1.52, indicating a 52.0% increase in’risk’, meaning reaching 52.0% more people from the priority population than from the general population. The practicality rating for this combination is high because both CDs are physical locations that can be visited. Alternatively, when using the’AND’ option, indicating the overlap between the CDs, targeting men who both regularly visit bars and beerhalls and have poor mental health would target only 7.5% of the general population and reach 14.8% of the priority population. The associated RR is 1.46, and the odds are 2.77 times higher for individuals in the priority population to have both CDs compared to those in the general population. The practicality rating for this combination is average, as it can be more challenging to target men with poor mental health. Targeting three CDs, for example: 1) residence on agricultural estate compounds; 2) negative social capital; and 3) poor mental health, could reach 57.1% of the priority population by targeting 29.9% of the general population.

In general, CD reach is lower in women than in men. For example, when targeting women on agricultural estate compounds AND/OR those who frequently visit bars and beerhalls, 15.8% of the general population would be targeted, and 20.9% of the priority population could be reached (Table 2). The’AND’ approach would have minimal reach, with all percentages below 1.0%.

A heatmap visualising RRs for the AND/OR approach for combinations of two CDs is shown in Figure 1. For both men and women, all high-value RRs involve the CD regular visits to bars or beerhalls. RRs generally are lower for women than for men, indicating that when targeting the general male population, a higher percentage of the priority population could be reached with HIV prevention programmes.

**Figure 1:**
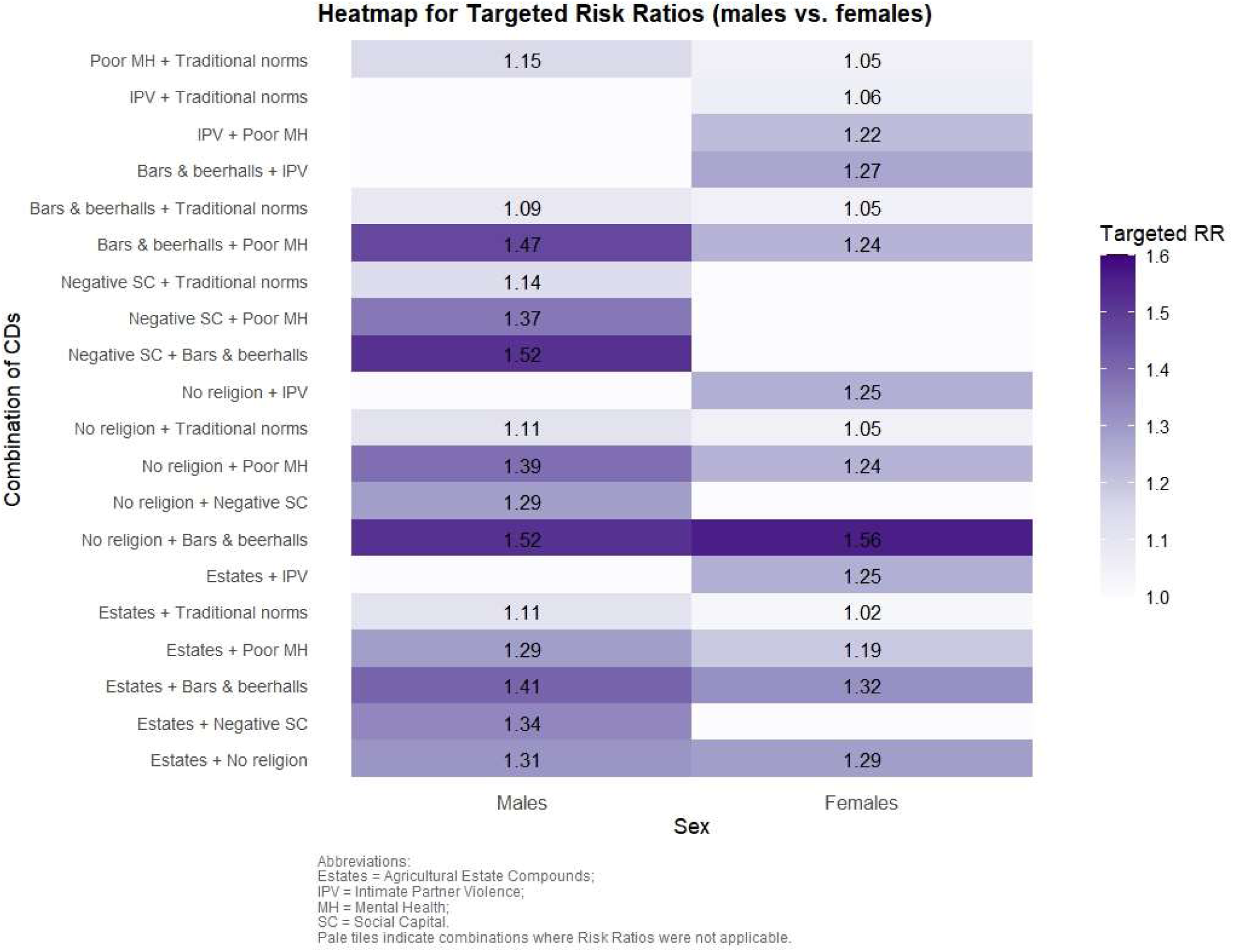
Heatmap of targeted Risk Ratios (RRs) for combinations of two Circumstantial Determinants (CDs) for the’AND/OR’ approach (targeting one or both CDs, with overlap considered) comparing males and females using data from east Zimbabwe.

### ROC curve algorithm identification of the priority population for HIV prevention

In men, the first AUC-based ROC curve (algorithm 1) demonstrated modest discriminatory ability (Figure 2A&B). The AUC in step 1 was 0.62 for the training dataset and 0.60 for the test dataset with the inclusion of regular visits to bars or beerhalls. Following this, the AUC increased gradually as more CDs were added, reaching a maximum of 0.63 in both datasets. Sensitivity increased stepwise from approximately 48.5% to 95.5% in the final step. Specificity declined from 74.9% to 15.9% in the training dataset and to 18.6% in the test dataset. Accuracy was highest at the initial step, at 70.4% in the training dataset and 67.5% in the test dataset and decreased in subsequent steps (Bagnay et al., 2026, Section 8.1). Patterns were consistent across both AUC-based algorithms, although the order of CDs differed in algorithm 2. Results from the sensitivity-based algorithm showed high sensitivity (over 86.0%) but considerably lower specificity, lower LR+, and AUCs that remained below 0.54. See Bagnay et al. (2026) for more information on Positive Predictive Values (PPV) and Negative Predictive Values (NPV).

**Figure 2:**
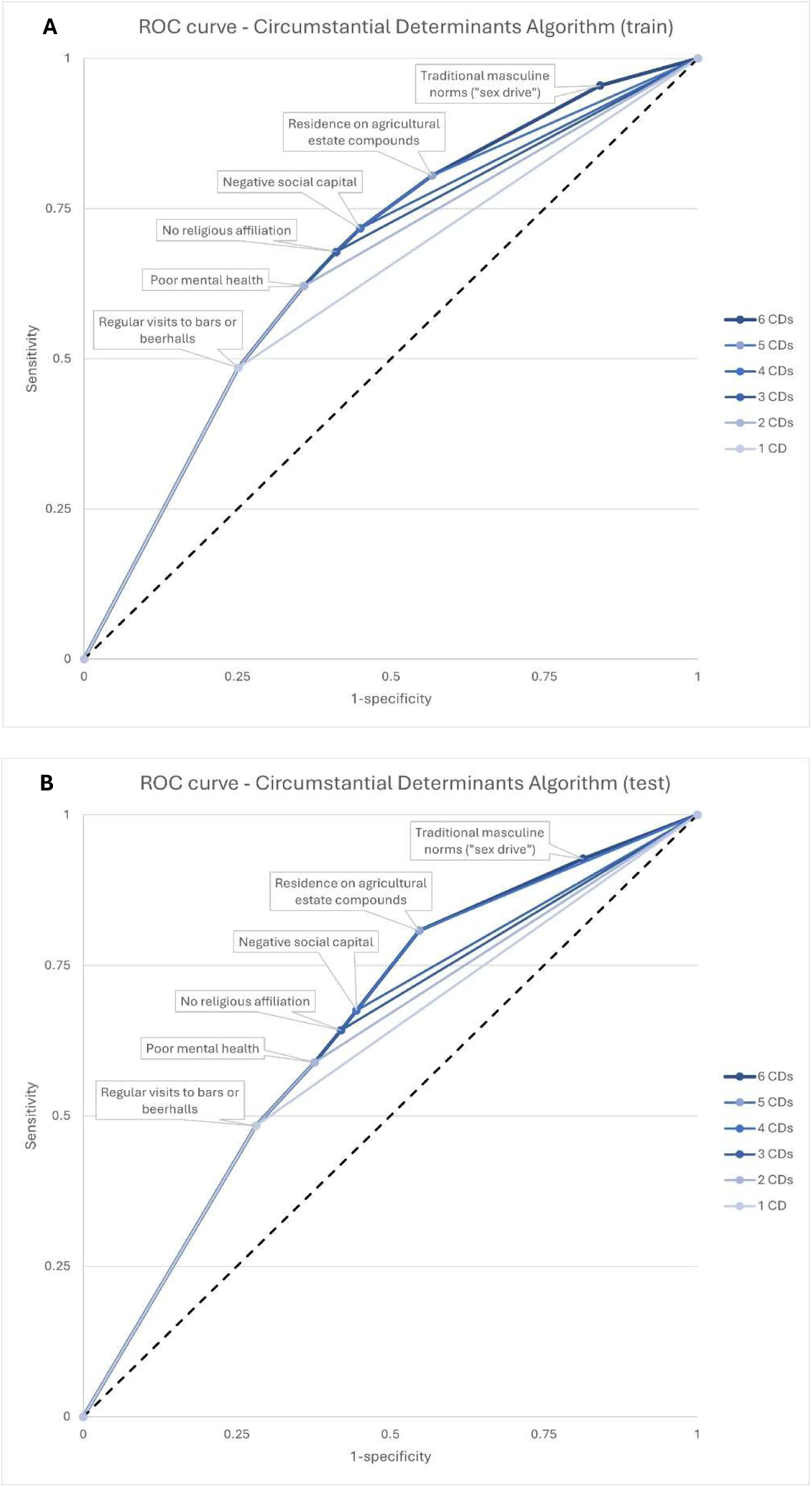
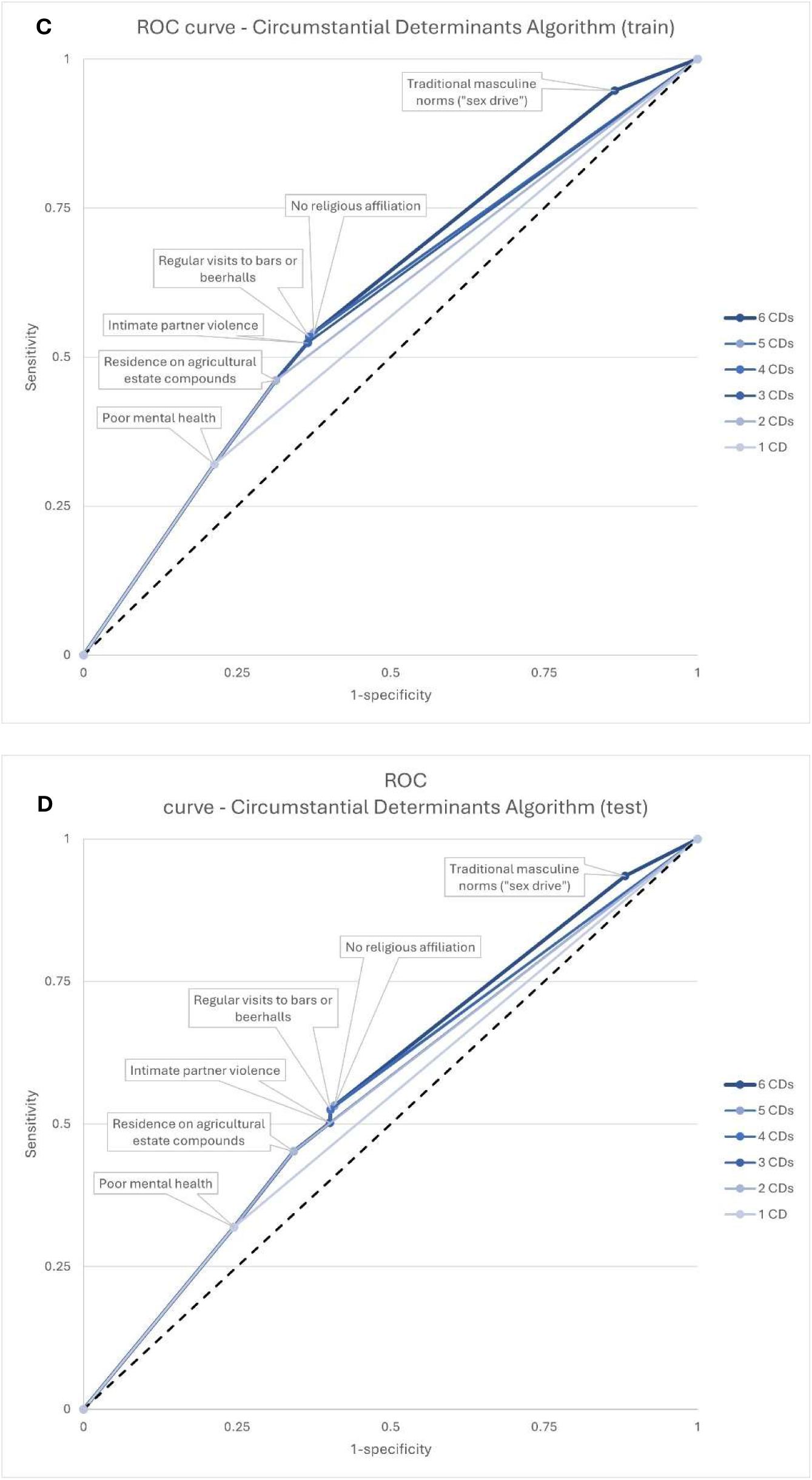
Receiver Operating Characteristic curves (algorithm 1) based on AUC with sensitivity plotted against (1-specificity) for HIV-negative males (A&B) or females (C&D), using data from the Manicaland 2018-2019 survey. Curves A and C display the results of the training datasets, while B and D show the outcomes of the test datasets.

In females, algorithm 1 again showed modest discriminatory ability (Figure 2C&D). The AUC in step 1 was 0.55 for females in the training dataset and 0.54 in the test dataset with the inclusion of experiencing poor mental health. As more CDs were added, sensitivity increased while specificity declined. The algorithm’s performance improved, but the AUC remained below 0.60. The highest accuracy in both datasets was achieved in the first step, with 68.5% in the training dataset and 65.1% in the test dataset (Bagnay et al., 2026, Section 8.1). Patterns in the other algorithms showed similar results, although the order of CDs varied. Additionally, results from the sensitivity-based algorithm indicated high sensitivity but with a specificity below 20%. The AUC was just above 0.50, peaking at 0.54.

In algorithm 1, for both males and females, gains in sensitivity in the training algorithm became smaller than the losses in specificity after step 4. In the test algorithms, this shift happened after step 5 in males and after step 2 in females.

### Comparison of HIV prevention condom cascades for priority population sub-groups reached by targeting different circumstantial determinants

Figure 3 shows selected examples of how male condom HIV prevention cascades – and therefore HIV prevention programme requirements – can differ between the sub-groups of the priority population for HIV prevention that could be reached by targeting different CDs.

Among men who regularly visit bars or beerhalls (Figure 3A), 74.3% were motivated to use condoms. Of these, 62.8% (or 46.7% of all men from the priority population for this CD) had access to male condoms, and 67.5% of those who were both motivated and had access (or 31.5% of all men from the priority population for this CD) reported effectively using condoms. Among men reporting poor mental health (Figure 3B), 76.8% were motivated to use condoms. Of these, 44.8% (or 34.4% of all men from the priority population for this CD) reported having access to condoms. Finally, 63.5% (or 21.9% of all men from the priority population for this CD) of those who were motivated and had access also effectively used male condoms.

**Figure 3:**
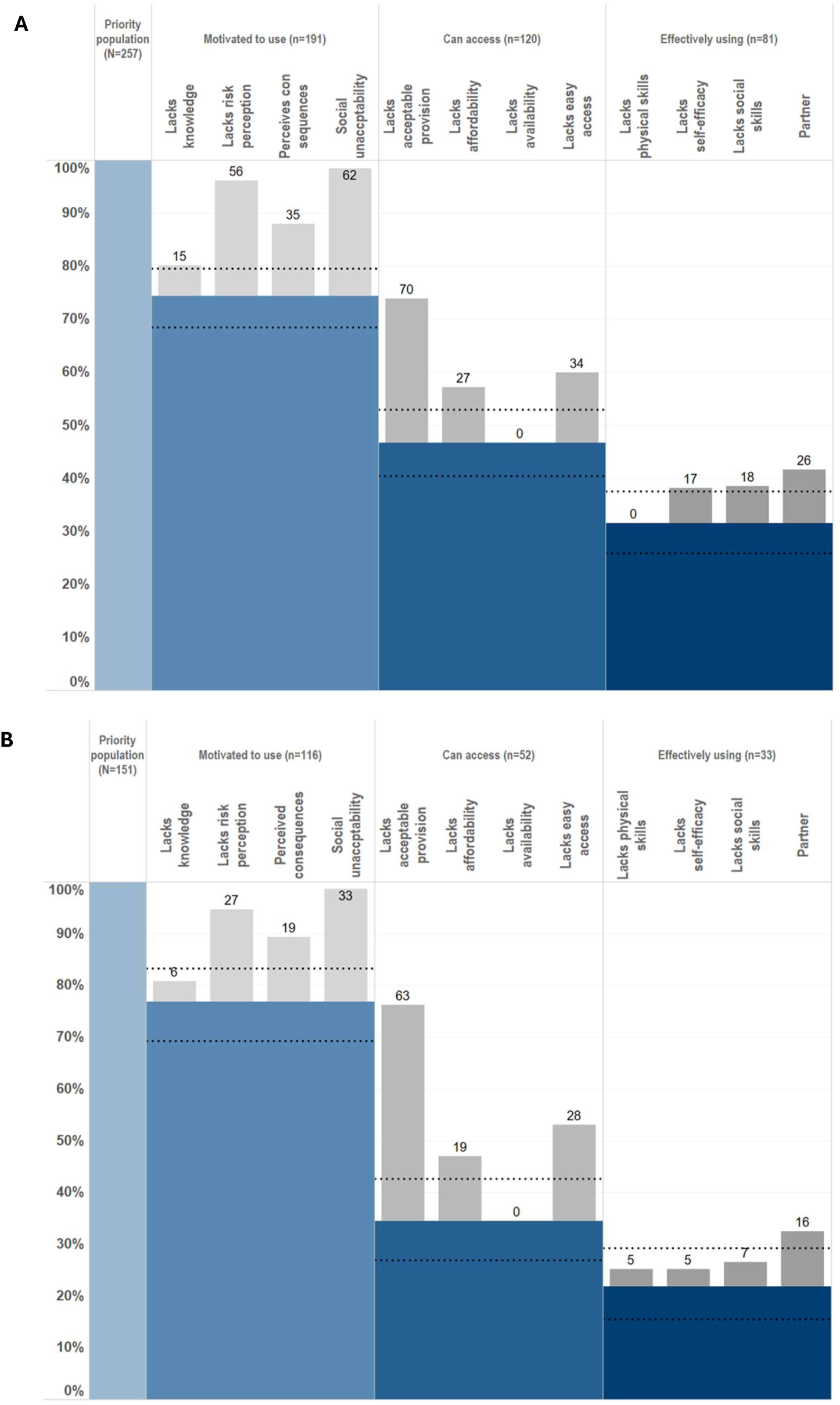
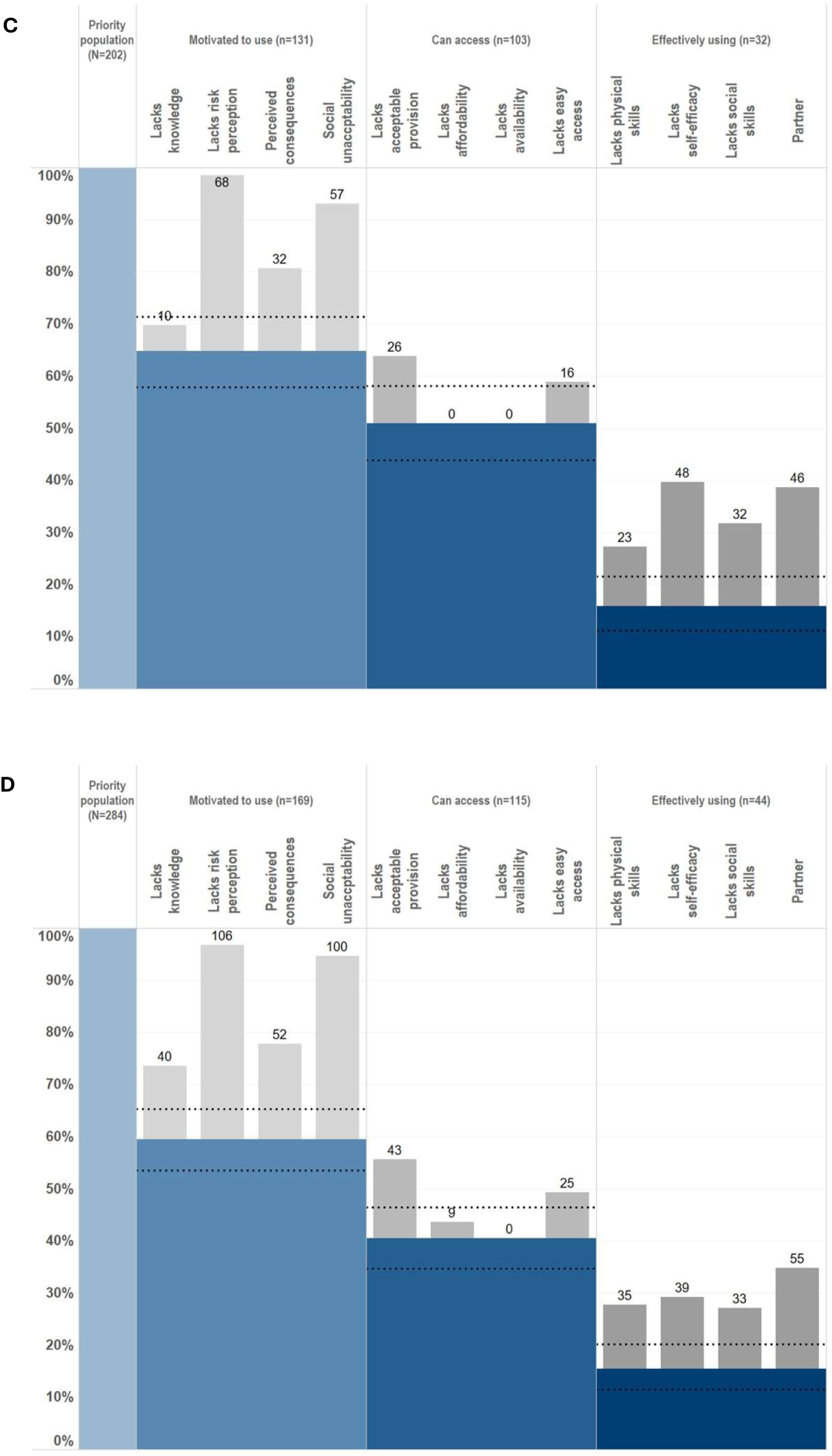
HIV prevention condom cascades for males (A&B) and females (C&D) from Manicaland, Zimbabwe, for selected circumstantial determinants. For males: (A) regularly visiting bars or beerhalls; (B) poor mental health. For females: (C) residence on agricultural estate compounds; (D) poor mental health. The first bar illustrates the’priority population’ per CD included within each HPCC. The dotted lines represent the 95%CI associated with each of the main bars.

Among women living on agricultural estate compounds (Figure 3C), 64.9% were motivated to use condoms. 78.6% (or 51.0% of all women from the priority population for this CD) of motivated women also reported having access to male condoms. Of those who were both motivated and had access, 31.1% (or 15.8% of all women from the priority population for this CD) effectively used condoms. Among women who reported poor mental health (Figure 3D), 59.5% were motivated to use male condoms. Of these, 68.0% (or 40.5% of all women from the priority population for this CD) reported having access to condoms. Among women both motivated and with access to condoms, 38.3% (or 15.5% of all women from the priority population for this CD) were effectively using condoms.

Differences in the main cascade barriers to use of male condoms for HIV prevention were found for both men and women (Bagnay et al., 2026, Section 9). Men with poor mental health were less likely to report having access to male condoms compared to those who regularly visit bars or beerhalls [OR: 0.37; 95% CI 0.20-0.68]. However, for men, the explanatory factors (i.e. cascade sub-bars) followed a similar pattern for both circumstantial determinants, with no statistically significant differences observed. For women, those experiencing poor mental health were more likely to report effective use of male condoms compared to those residing in agricultural estate compounds [1.94; 1.19-3.18]. Several statistically significant differences in cascade sub-bars were found (Table 9.2b, Section 9). Among women in the priority population, those experiencing poor mental health had four times higher odds of low knowledge about HIV, which acts as a barrier to motivation to use condoms, compared to women living on agricultural estate compounds. Additionally, among women who were motivated and had access, those experiencing poor mental health had over three times higher odds of lacking practical skills and facing partner-related barriers to effective condom use compared to women residing on agricultural estate compounds. Conversely, these women had lower odds of reporting self-efficacy as a barrier to effectively using male condoms compared to women living on agricultural estate compounds.

## Discussion

We investigated the potential utility of a targeted Circumstantial Determinants approach to reaching people in the general population at higher risk of acquiring HIV infection with HIV prevention and testing services. The idea being that this approach would build from and extend the scope of existing strategies that target social contexts – most notably early programmes with men and women at bars and beerhalls (Fritz et al., 2011) and the PLACE strategy (Weir et al., 2003).

We found that, in Manicaland, east Zimbabwe, incrementally targeting more circumstantial determinants could significantly expand the reach of such programmes into the priority population for use of HIV prevention methods. For instance, we observed that targeting men at bars and beerhalls could reach 48.5% of the priority population in need of HIV prevention. This reach could potentially increase to 77.1% if those with poor mental health, no religious affiliation, negative social capital, or living on an agricultural estate were also included. The’cost’ of this considerable increase in coverage would be that the percentage targeted of those who are at lower risk of HIV infection would rise from 25.1% to 53.7%. However, this additional inclusion of lower-risk individuals would nonetheless remain substantially smaller than in an untargeted approach. There will always be a trade-off between sensitivity and specificity where the implications of increasing coverage for those at priority are weighed against the implications of including more of those with no risk. The optimum strategy will depend on the cost of the intervention being promoted.

For women, we found that experiencing poor mental health was the most impactful circumstantial determinant for increasing the AUC, and targeting these women could reach 32.0% of the priority population for HIV prevention and 21.3% of lower-risk women. If, for example, women living on agricultural estates, experiencing intimate partner violence, attending bars and beerhalls, or having no religious affiliation were also targeted, coverage could increase to 54.1%. In this scenario, 37.5% of women at lower risk of HIV infection would also be targeted. In both cases, the algorithm optimised using the training dataset produced very similar results when applied to the test dataset.

These findings reflect two key points: first, that for several CDs, the relative risk that individuals in the general population exposed to these determinants are part of the priority population is significantly greater than one; and second, that targeting individuals exposed to any two different CDs often results in reaching different segments of the priority population. Examples of Venn diagrams in Bagnay et al. (2026), Section 7, illustrate this.

Interestingly, we also found differences in the gaps between the main bars in the HIV prevention cascades for male condoms among men and women in the priority population exposed to various CDs. For example, men experiencing poor mental health were less likely to have access to condoms than those who frequently visit bars and beerhalls, while women from the same group were more likely to use condoms effectively than women living in agricultural estate compounds. Men and women in the priority population reporting exposure to different CDs also differed in their sources of information about HIV prevention (Bagnay et al., 2026, Section 10). These findings suggest that varying the emphases on intervention activities and messaging aimed at addressing barriers to motivation, access, and capacity to use HIV prevention methods, respectively, could be deployed when targeting different CDs to increase the overall impact of HIV prevention programmes.

The strengths of this study include the availability of linked data on a wide range of locally relevant CDs, sexual risk-behaviours for HIV acquisition, and barriers to condom use for HIV prevention for the same population. The data were drawn from a good-sized general population sample of HIV-negative individuals across all main socio-economic strata, except mining areas, found in Zimbabwe’s Manicaland province, which should enhance the local representativeness of the study findings. However, the strength of association between exposure to any given CD and HIV risk will differ between populations and over time, as will the extent of overlap between exposures to different CDs. Therefore, further research will be needed in different locations and at different time points to assess the wider generalisability of the study findings - regarding both the broader utility of the overall Circumstantial Determinants approach and the individual CDs required to be included in any given setting. Informal confidential voting interview methods (Gregson et al., 2002) were used in the data collection to reduce social desirability bias in reports on sexual behaviour. However, some under-reporting will still be present in the data and may have affected the ROC-curve results. Other limitations include the study’s cross-sectional nature – making it difficult to draw causal inferences – and the inclusion of only male condoms in the HIV prevention cascade analysis.

Applying the Circumstantial Determinants approach in practice will require developing, locally selecting, and adapting sets of non-stigmatising and non-discriminatory strategies to reach people exposed to a number of contrasting circumstances. For some CDs – for example, in Manicaland, targeting men and women living on agricultural estate compounds – this could be relatively straightforward. However, for other circumstantial determinants (e.g. poor mental health, intimate partner violence, and no religious affiliation), it will be more challenging, and it may only be possible to reach subsets of individuals exposed to these circumstances. For example, for mental health, those with severe conditions could be reached by targeting individuals attending routine health services. Reaching individuals with less severe conditions would be more challenging but could perhaps be done through collaborations with community-based mental health programmes such as the Friendship Bench (Chibanda, 2017). To develop a broadly applicable approach, identification of CDs and strategies to reach those within priority populations will need to be explored locally until generalisable patterns emerge. Beyond targeting, the circumstantial determinants identified in this study may themselves be sites for structural interventions. Thus, addressing these underlying circumstances could reduce HIV transmission while yielding wider health benefits, with knock-on effects on other diseases driven by similar circumstances.

Activities to address many of the common barriers in HIV prevention cascades could be implemented productively with all individuals reached through CDs without further screening – i.e. regardless of recent or current risk behaviours – since everyone in populations with generalised HIV epidemics is at some degree of risk of acquiring or transmitting the infection. Such activities could include those to improve knowledge about HIV and the accuracy of HIV risk perception (Schaefer et al., 2019) and to address misunderstandings about the negative consequences of using HIV prevention methods or barriers to access these methods. However, when there is a strong focus on promoting HIV prevention methods for use by uninfected individuals, including PrEP (Pike et al., 2025) and VMMC (Mavhu et al., 2021), HIV testing will be needed and, for those with seronegative results, further screening for HIV risk may be necessary. In such situations, under-reporting by individuals in the priority population for HIV prevention might be reduced by including questions about’seasons of risk’ (Pickles et al., 2025) and cumulative life-course exposures (Vineis & Barouki, 2022) in the screening tools used to assess for current and future HIV risk. More generally, when screening is done in circumstantial determinant contexts widely recognised as being associated with HIV risk, social desirability bias may be a smaller barrier to accurate reporting of sexual risk behaviours.

Models of HIV prevention cost-effectiveness often assume that priority populations are reached (Jamieson et al., 2020; Wu et al., 2024). Our results show how well priority populations could be covered and could be used to better parameterise such analyses.

The results of this exploratory study are encouraging and suggest that the Circumstantial Determinants approach has strong potential to improve the targeting of HIV prevention and testing services. Whilst the focus of this paper was on HIV prevention amongst currently uninfected adults at risk of newly acquiring HIV infection, it is plausible that, where status-neutral HIV testing is done, the same approach could also be effective in reaching newly infected people living with HIV for enrolment in national ART programmes. Next steps should include refining the overall approach through research in other settings, conducting feasibility studies of detailed strategies to maximise programme reach in people exposed to different circumstantial determinants, mathematical modelling of the population-level impact of implementing the approach under alternative scenarios, and cost-effectiveness studies.

## Data Availability

Given the sensitive nature of the data collected, including details about HIV status, treatment and sexual risk behaviour, the Manicaland Centre for Public Health does not make full analysis datasets publicly available. Nevertheless, summary datasets encompassing household and background sociodemographic individual questionnaire data from rounds 1 to 9 (1998 to 2023) are available for download via the Manicaland Centre website. Additionally, summary data on HIV incidence and mortality from rounds 1 to 6 (1998 to 2013), developed in collaboration with the ALPHA Network, are available through the DataFirst Repository. For other quantitative data used in the Manicaland Centre for Public Health's analyses, interested parties can request information by completing a data access request form.

http://www.manicalandhivproject.org/data-access.html

https://www.datafirst.uct.ac.za/dataportal/index.php/catalog/ALPHA/about

http://www.manicalandhivproject.org/data-access.html

## Notes

### Competing Interest Statement

Simon Gregson declares shareholdings in pharmaceutical companies (GlaxoSmithKline and AstraZeneca). All other authors have no conflicts of interest to declare.

### Author Declarations

Ethical approval was obtained from the Imperial College Research Ethics Committee (17IC4160) and the Medical Research Council of Zimbabwe (MRCZ/A/2243). All participants in this study provided signed informed consent. For participants under 18, written informed consent was obtained from a parent or guardian, and informed assent was obtained from the adolescent. Data has been anonymised to ensure participants cannot be identified.

### Funding

- Gates Foundation: INV-023210
- Medical Research Council: MR/X020258/1

